# Allostatic Load in Adults with Congenital Heart Disease: A Multi-Cohort Analysis of the All of Us Research Program

**DOI:** 10.64898/2026.05.19.26353630

**Authors:** Michael T. M. Finn, Stefania Soria, Gruschen R. Veldtman

**Affiliations:** Department of Pediatrics and Human Development, Michigan State University College of Human Medicine, Grand Rapids, MI; Betz Congenital Heart Center, Corewell Health Helen DeVos Children’s Hospital, Grand Rapids, MI

**Keywords:** congenital heart disease, allostatic load, multimorbidity, mental health, *All of Us*

## Abstract

**Background:** Adults with congenital heart disease (CHD) are a growing population and face unique challenges as they age. Unlike acquired diseases that disrupt a previously healthy baseline, CHD is developmentally embedded. Allostatic load, the multi-system biological “wear and tear” exacted by the continuous cost of coping, offers a framework for indexing this lifelong psychophysiological stress.

**Methods:** We analyzed 14,469 adults from the *All of Us* Research Program: non-syndromic CHD (*n* = 6,810), acquired heart disease (AHD; *n* = 2,264), non-cardiac chronic illness (*n* = 4,331), and a general population comparison cohort (GP; *n* = 1,064). Using a standardized operationalization, allostatic load was scored across five biomarker domains (AL5, range 0-5). A pre-specified primary test compared adjusted AL5 between CHD and GP. Exploratory analyses examined clinical predictor of this gap and whether baseline subjective health predicted prospective AL5 change, utilizing strictly matched biomarkers across timepoints to prevent substitution artifacts.

**Results:** Adults with CHD carried significantly higher allostatic load than the general population comparison cohort (adjusted difference +0.30 AL5 units, 95% CI 0.24-0.37, *p* < .001). Cumulative comorbidity and cardiac medication burden explained most of this gap. Congenital anatomical complexity did not independently predict this burden. In a prospective subsample (*n* = 8,031, mean follow-up 2.7 years), worse baseline mental health predicted increases in allostatic load over time in CHD. Baseline physical health showed no such prospective association. The general population and acquired heart disease cohorts demonstrated the inverse dissociation: subjective physical health predicted these longitudinal physiological changes.

**Conclusions:** Adults with CHD carry an elevated allostatic burden dictated by the cumulative cost of acquired medical and treatment intensity. The original congenital anatomy does not predict this accumulation. Furthermore, subjective mental health prospectively tracks future increases in allostatic load in CHD. This dissociation is absent in adult-onset acquired heart disease, suggesting that the mental aspects of coping with CHD may impact outcomes above and beyond those with acquired heart disease. These findings position psychological care as a potentially physiologically consequential intervention.

## Introduction

Adults with congenital heart disease now outnumber children with the condition.^1,2^ Survival has revealed new clinical realities that adults with congenital heart disease may face. Many of these patients live in long-term medical stability, but they nonetheless carry chronic fatigue, exercise limitation, and psychological distress that residual anatomy and physiology cannot fully explain.^3-5^ In adult-onset acquired heart disease, such symptoms typically emerge when an illness disrupts a previously healthy physiological baseline. Congenital heart disease is fundamentally different. Because the condition is developmentally embedded, it forms the patient’s physiological baseline from the beginning of life, rather than the typical perspectives of recovery being a return to a prior healthy state.^6^ Their cardiovascular and autonomic nervous systems have co-developed under structural constraint since fetal life. Capturing the cumulative cost of this lifelong adaptation requires a measure that spans multiple organ systems, rather than any single snapshot of cardiac severity.

Allostatic load is one such measure.^7,8^ Originally conceptualized by McEwen, the construct frames chronic physiological wear as the active “cost of coping.”^7^ It posits that the repeated or sustained engagement of stress-response systems to maintain systemic stability leaves a measurable biomarker footprint across the multiple physiological systems.^8,9^ For adults with CHD, this framework is particularly apt. The systemic strain they carry is rarely an episodic reaction to an acute stressor; rather, it may be the continuous physiological price of holding a constrained physiological baseline together against the ordinary demands of daily life.

Translating this concept into large-scale clinical databases requires an objective and practical approach. To achieve this, we utilized a standardized allostatic load protocol designed specifically for electronic health record environments,^10^ anchored to the five-biomarker index identified by a recent multi-cohort individual participant data meta-analysis as the most reliably associated with multi-system dysregulation and mortality.^11^ The result is a straightforward approach where one calculates a cumulative load score based on the count out of five biomarkers exceeding high-risk population thresholds. This provides a reproducible index of cumulative wear from chronic stress.

To date, no study has examined allostatic load adults with CHD, despite clear evidence of elevated psychological disorders, stress, and early life adversity in this population.^4^ We hypothesized that adults with CHD would show a higher allostatic load than the general population. This provides an initial test of whether lifelong cardiac survivorship leaves a measurable systemic footprint. We then explored what factors predicted any established gap between CHD and general population in allostatic load, whether subjective health predicts changes on this measure over time, and which individual biomarker domains tend to shift most over time. We also included two additional matched cohorts for descriptive context: acquired heart disease (AHD) and non-cardiac chronic illness (NCCI).

## Method

### Data source

Data were from the All of Us Research Program controlled-tier release.^12^

### Cohort construction

We constructed four cohorts: the target congenital heart disease cohort, an acquired heart disease cohort, a non-cardiac chronic illness cohort, and a general population cohort. See Figure 1 for a diagram of the matching and flow of data availability to the analytic samples.

**Figure 1.**
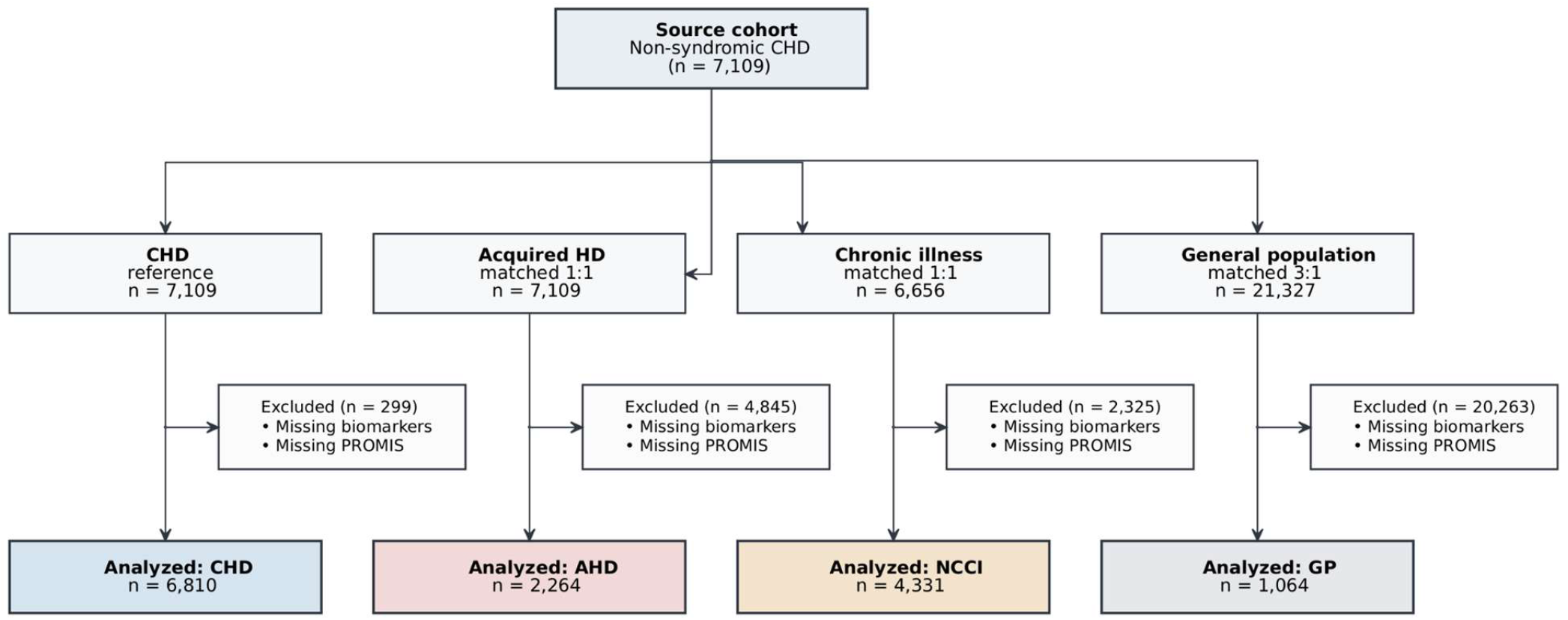
Cohort Construction and Matching Flowchart. **Legend:** Flow diagram detailing the derivation of the analytic sample from the *All of Us* Research Program controlled-tier dataset. The target congenital heart disease (CHD) cohort was restricted to adults with definitive structural congenital codes. Multisystem syndromic/chromosomal conditions and participants missing required baseline biomarkers or PROMIS surveys were excluded. The three comparison cohorts, Acquired Heart Disease (AHD), Non-Cardiac Chronic Illness (NCCI), and the General Population (GP), were initially drawn using exact-sex and nearest-neighbor age matching (+/-2-year caliper) prior to complete-case exclusions. The final analytic sample comprised 14,469 adults for cross-sectional evaluation, with a subset of 8,031 adults retaining paired longitudinal biomarkers for prospective analyses (mean follow-up 2.7 years).

#### Congenital heart disease (CHD; n = 6,810)

Adults with at least one ICD-9/ICD-10 code for a congenital structural heart defect, drawn from ICD-9 745-747 and 759.3 and ICD-10 Q20-Q26 and Q89.3. Participants with co-occurring chromosomal or syndromic diagnoses were excluded. Anatomical complexity (Simple, Moderate, Complex) was assigned.^13^ Full code lists, exclusion logic, and tiering rules are in Supplementary Section A.

#### Acquired heart disease (AHD; n = 2,264)

Adults with ischemic heart disease, heart failure, cardiomyopathy, arrhythmia, or non-congenital valvular disease and no CHD codes.

#### Non-cardiac chronic illness (NCCI; n = 4,331)

Adults with at least one Charlson-level chronic condition (malignancy, diabetes, chronic kidney disease, chronic obstructive pulmonary disease, chronic liver disease) and no cardiac codes.

#### General population (GP; n = 1,064)

Adults with none of the above and no codes indicating prior cardiac surgery, transplant, or otherwise present cardiac disease.

Comparison pools were initially drawn using exact matching on biological sex and nearest-neighbor matching on age within a ±2-year caliper (3:1 for GP; 1:1 for AHD and NCCI) to the CHD cohort. However, the subsequent strict requirement for complete psychometric and biomarker data resulted in asymmetric missing-data attrition, yielding final analytic cohorts with residual demographic imbalances (Supplementary Table S1). All regression models utilized covariate adjustment to account for these residual demographic differences.

### Measures

#### Allostatic load (AL5)

Cumulative physiological wear was quantified using a five-component allostatic load score (AL5, range 0-5) based on the CHORDS (Community Health Oriented Resilience Data Science) protocol.^10^ Developed specifically to operationalize allostatic load within large-scale electronic health record environments like the All of Us Research Program, this standardized framework assigns one point for each biomarker exceeding high-risk clinical thresholds across five domains: inflammation, glycemia, lipids, autonomic tone, and anthropometry (Table 1). We made one modification to the CHORDS protocol given the relative lack of CRP in our sample. White blood cell count (WBC) was deployed as a pre-specified fallback proxy for the inflammatory domain when C-reactive protein (CRP) was unavailable.

**Table 1.** Allostatic Load Score (AL5): Domains, Biomarkers, and High-Risk Thresholds.

| Domain | Biomarker | High-risk threshold | Clinical equivalent |
| --- | --- | --- | --- |
| **Inflammation** | C-reactive protein (CRP)* | ≥ cohort 75th percentile | $\geq 3$ mg/L |
| **Glycemia** | Hemoglobin A1c | ≥ cohort 75th percentile | $\geq 5.7\%$ |
| **Lipids** | HDL cholesterol | ≤ cohort 25th percentile | $< 40$ mg/dL (men) / $< 50$ mg/dL (women) |
| **Autonomic tone** | Resting heart rate | ≥ cohort 75th percentile | $\geq 90$ bpm |
| **Anthropometry** | Waist-to-hip ratio | ≥ cohort 75th percentile | $\geq 0.90$ (men) / $\geq 0.85$ (women) |

*Note: White blood cell count (WBC, elevated at ≥ cohort 75th percentile) served as a fallback for missing CRP to maximize cohort retention in cross-sectional and baseline models; all longitudinal analyses strictly required paired CRP*
1 *measurements to prevent substitution artifacts. $AL5$ = sum* 2 *across the five domains (range 0–5).*

To capture both static burden and dynamic physiological weathering, we operationalized two discrete AL5 values per participant: a survey-anchored AL5 (derived from the biomarker panel temporally closest to the PROMIS survey completion) which served as the prospective baseline, and a most-recent AL5 which served as both the cross-sectional exposure and the longitudinal follow-up.

Crucially, to ensure that estimated physiological trajectories reflected genuine biological weathering rather than measurement artifacts, all longitudinal change and component-level drift analyses were strictly constrained to matched biomarkers. Full OMOP concept IDs, unit harmonization protocols, and quality control logic are detailed in Supplementary Section B.

#### Subjective health

The PROMIS Global Health short form^14^ provided Mental and Physical heath rating T-scores at a single baseline timepoint.

#### Covariates

The Charlson Comorbidity Index^15^ summarized non-cardiac chronic disease. Acquired cardiac disease burden was a count (0-7) of distinct acquired cardiac domains (heart failure, atrial arrhythmia, ventricular arrhythmia, coronary artery disease, pulmonary hypertension, endocarditis, aortic disease). Cardiac medication burden was a count (0-3) of cardiac drug classes (lipid-lowering, antihypertensive, antidiabetic) over the prior five years. Healthcare utilization, mental health diagnosis, and psychosocial covariates (smoking, food insecurity, financial barriers to care, social support, loneliness, perceived stress, health literacy) were also extracted; full definitions are in Supplementary Section C.

## Statistical analysis

### Primary confirmatory test

The CHD-GP AL5 difference was tested using demographics-adjusted estimated marginal means with Dunnett-adjusted contrasts. The CHD-vs-GP contrast was the pre-specified primary test.

### Exploratory analyses

We then pursued three exploratory questions. 1) *What explains the gap?* Six regression blocks added covariates in turn (demographics → Charlson index → acquired cardiac burden → cardiac medications → utilization and psychosocial factors → PROMIS), tracking how the CHD-GP gap changed at each step. Generalized variance inflation factors confirmed that this attenuation reflected substantive overlap of explanatory variables, not collinear shrinkage. Within the CHD cohort, we used Type III ANOVA to identify which clinical features most strongly predicted AL5. As a complementary descriptive analysis, we used Shapley variance decomposition^16^ to apportion within-cohort AL5 variance across four predictor groups and test the degree of variance accounted for independent of their order of entry into hierarchical regression models (medical burden; demographics; subjective health; social/behavioral).

1. *Does subjective health predict biomarker change?* In the prospective subsample (n = 8,031 participants with two complete biomarker panels separated by at least 6 months), we fit a linear interaction model predicting AL5 change from baseline PROMIS Mental and Physical T-scores by cohort, adjusted for demographics, Charlson index, baseline AL5, and follow-up interval. Because PROMIS was assessed only once at baseline in the All of Us protocol, we conducted sensitivity analyses examining whether the observed associations were stable across baseline AL5 levels and follow-up intervals (that is, patterns that would not be expected under regression-to-mean or noise-dilution artifacts).
2. *Component-level drift over time*. We explored which AL5 components changed most across cohorts by fitting a logistic regression of biomarker worsening on cohort, adjusting for age, sex, race, Charlson index, and inter-observation interval. We did not utilized the WBC fallback for CRP in this analysis.

All analyses were conducted in R version 4.3 using the *tidyverse, lubridate, emmeans, car*, and *relaimpo* packages.

## Results

### Cohort construction

Cohort construction is shown in Figure 1. Of 7,109 adults meeting CHD inclusion criteria, 6,810 had complete biomarker and PROMIS data and entered analyses. Comparison cohorts after initial matching and subsequent complete-case exclusion comprised 2,264 AHD, 4,331 NCCI, and 1,064 GP participants. Due to differential missing-data attrition, residual demographic imbalances remained across the final analytic cohorts (Supplementary Table S1), requiring demographics adjustment in all models.

### Adults with CHD carry higher allostatic load than general population

The primary hypothesis was supported. Adults with CHD showed a demographics-adjusted AL5 of 0.90 (95% CI 0.88-0.93) compared with 0.60 (95% CI 0.54-0.66) in the general population comparison cohort, a difference of +0.30 AL5 units (*p* < .001; Figure 2). For context, the AHD cohort showed a larger adjusted difference relative to GP (+0.51) as did the NCCI cohort (+0.37). That AHD exceeded CHD on this index, despite CHD being lifelong, is worth a brief note. This pattern is consistent with AHD representing recent dysregulation against a previously intact baseline. CHD’s long-standing physiological adaptation may produce a biomarker snapshot that underestimates the underlying regulatory cost.

**Figure 2.**
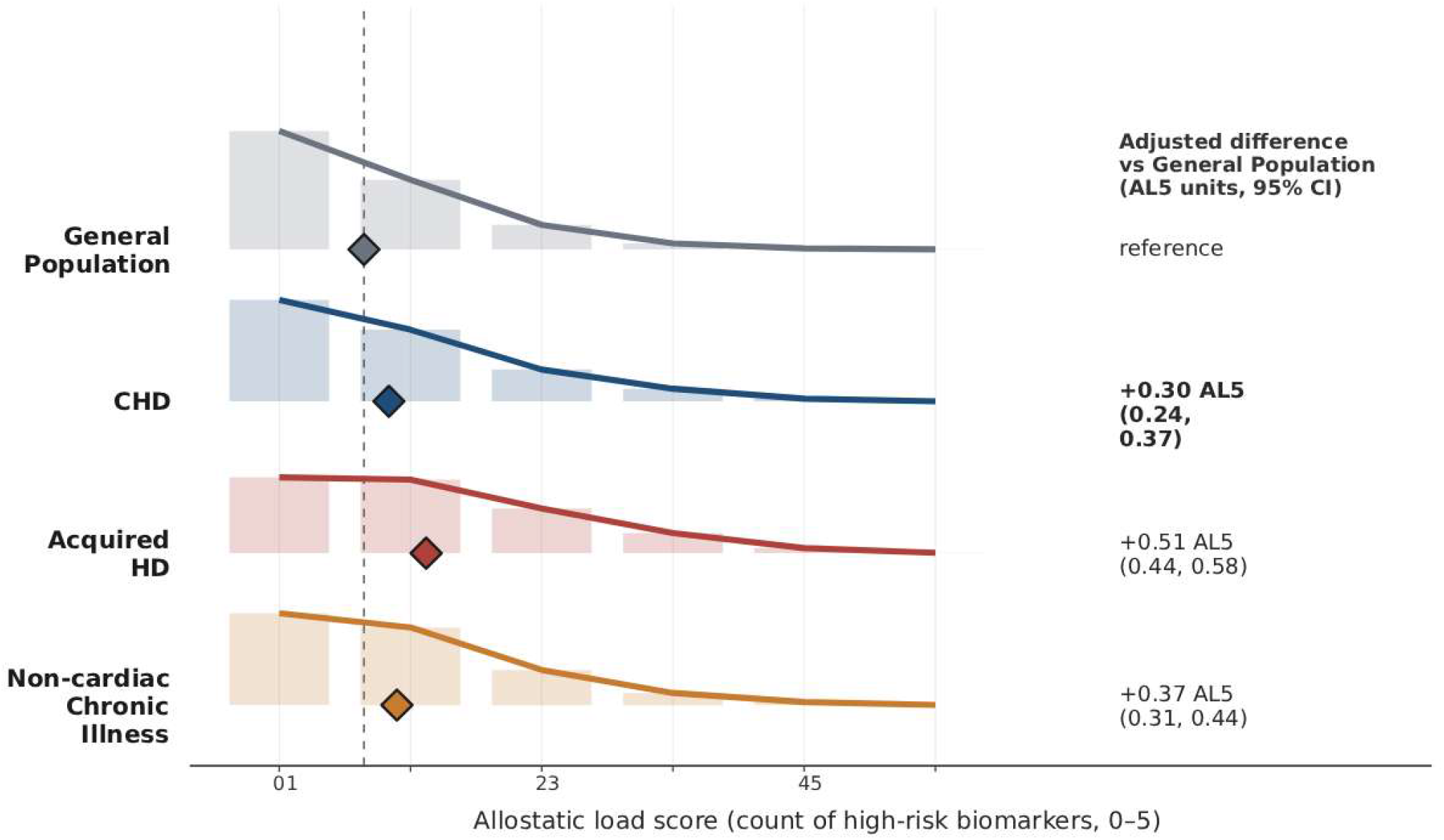
Elevated Allostatic Load in Adults with Congenital Heart Disease. **Legend:** Cross-sectional distribution of allostatic load (AL5; 0-5 score) across the four analyzed cohorts. AL5 scores are expressed relative to the matched general population (GP) reference zero-line. Half-violin plots demonstrate cohort-level density, overlaid with jittered points representing individual participant scores. Diamonds indicate the unadjusted cohort means. The right-hand column displays the fully adjusted difference (and 95% CI) for each clinical cohort relative to the GP comparison group, controlling for age, sex, and race. Adults with CHD carry a significantly elevated physiological burden (+0.30 AL5 units), though this gap is narrower than that observed in adult-onset acquired heart disease (+0.51 AL5 units).

#### What explains the gap: accumulated burden, not anatomy

Cumulative comorbidity and cardiac treatment burden explained most of the CHD-GP gap in allostatic load score (Table 2).Adding the Charlson comorbidity index alone reduced the gap by two-thirds. Adding acquired cardiac disease burden and cardiac medication count reduced it further, with the residual coefficient becoming negative. Once acquired medical burden is accounted for, CHD itself no longer carries an independent positive contribution to allostatic load. Variance inflation across the full model was modest (maximum 1.81), indicating tolerable collinearity effects.

**Table 2.** What Explains the CHD-GP Allostatic Load Gap.

| **Block** | **Covariates added** | **Residual gap (SD)** | **Cumulative % of gap** | **Model R2** | **ΔR2** | **ΔR2 p-value** |
| --- | --- | --- | --- | --- | --- | --- |
| **1** | Demographics only | +0.29 | 3% | 0.07 | N/A | N/A |
| **2** | + Charlson comorbidity index | +0.10 | 69% | 0.12 | +0.05 | < .001 |
| **3** | + Acquired cardiac burden | -0.06 | 119% | 0.13 | +0.01 | < .001 |
| **4** | + Cardiac medications | -0.15 | 148% | 0.17 | +0.04 | < .001 |
| **5** | + Healthcare utilization, MH, social factors | -0.16 | 152% | 0.18 | +0.01 | < .001 |
| **6** | + PROMIS Mental and Physical | -0.15 | 149% | 0.19 | +0.01 | < .001 |

*Note: Percentages expressed relative to the demographics-adjusted Block 1 gap of +0.29* *AL5 Units. Maximum generalized variance inflation factor in Block 6 was 1.83, well*
below conventional concern thresholds. The negative residual coefficient from Block 3 onward indicates that once accumulated medical and treatment burden are accounted for, CHD itself no longer carries an independent positive contribution to AL5. Incremental *p*-values derived from nested likelihood ratio tests.

Probing the predictors of allostatic load within the CHD cohort alone, a similar pattern held. Cardiac medication burden was the strongest single predictor (F = 236, *p* < .001), exceeding both Charlson index (F = 135) and age (F = 115). Anatomical complexity, the lesion category assigned at diagnosis, did not significantly predict AL5 after adjustment for acquired burden (F = 2.5, *p* = .08). PROMIS Mental and Physical scores each contributed unique variance (F = 19 and 13, both *p* < .001). Full results are in Supplementary Table S4.

The progressive decomposition above orders covariates by clinical priority, which means the share of variance attributed to any one block depends on what was entered before it. To complement that ordering-dependent view with an ordering-invariant one, we used Shapley variance decomposition, which averages each block of predictors across all possible entry orders. Within the CHD cohort, this confirmed the picture from the progressive analysis: medical and system burden was the dominant contributor, accounting for 57.4% of the explained AL5 variance (absolute R^2^ contribution = 0.11). Demographics represented the second-largest share at 33.1% (R^2^ = 0.07), while subjective health (6.2%, R^2^ = 0.01) and SDOH/health behaviors (3.4%, R^2^ = 0.01) contributed smaller proportions of the total explained variance (Supplementary Table S3). A sensitivity analysis using the more inclusive single-occurrence definition of mental health diagnosis (Supplementary Table S6) yielded substantively identical cohort coefficients in the full Block 5 model, with shifts of less than 0.01 SD across all three cohort contrasts. The decomposition persists over operationalization of mental health diagnoses.

### Mental health prospectively tracks biomarker change in CHD

In the prospective subsample (*n* = 8,031; mean follow-up = 2.68 years, *SD* = 1.35), baseline subjective health predicted subsequent AL5 change in a cohort-specific pattern (Table 3). In CHD, baseline mental health exhibited a significant prospective association with the rate of biological weathering: higher Mental T-scores predicted attenuated allostatic load accumulation over follow-up (slope = -0.03 per 1-SD higher score; 95% *CI*, -0.043 to -0.014; *p* < .001), whereas worse baseline mental health predicted accelerated biomarker worsening.

**Table 3.** Baseline PROMIS Predicting AL5 Change Over Follow-Up, by Cohort.

| Cohort | Mental Health slope<br>(95% CI) | p | Physical Health slope<br>(95% CI) | p |
| --- | --- | --- | --- | --- |
| General population | +0.02 (-0.02, +0.05) | .42 | **-0.04 (-0.08, -0.01)** | **.014** |
| CHD | **-0.03 (-0.04, -0.01)** | **< .001** | +0.00 (-0.01, +0.02) | .72 |
| Acquired heart disease | -0.02 (-0.05, +0.00) | .10 | **-0.03 (-0.05, -0.01)** | **.011** |
| Non-cardiac chronic illness | **-0.03 (-0.05, -0.02)** | **< .001** | +0.00 (-0.01, +0.02) | .66 |

Note: Slopes are AL5 change per 1-SD higher baseline PROMIS T-score (negative = better baseline subjective health predicts less worsening). Adjusted for age, sex, race, Charlson index, baseline AL5, and inter-observation interval. Bold indicates significant cohort-specific slopes. Mean follow-up: 2.7 years.

Baseline physical health scores in CHD showed no such prospective association (*p* = .72). The general population and acquired heart disease cohorts demonstrated the inverse dissociation: subjective physical health, rather than mental health, prospectively predicted allostatic load change in those groups (Table 5). The non-cardiac chronic illness cohort (NCCI) followed the CHD pattern, with mental health rather than physical health predicting prospective physiological change.

PROMIS was assessed only at baseline, so we conducted two sensitivity checks to examine a genuine forward-effect interpretation over regression-to-mean or measurement noise (Supplementary Table S5). In CHD, the Mental → ΔAL5 slope was essentially uniform across baseline AL5 levels (-0.039, -0.042, -0.044 at -1 SD, mean, and +1 SD baseline AL5), bypassing the attenuated-at-high-baseline pattern expected under strong regression to mean. The slope also strengthened slightly with longer follow-up (-0.037, -0.041, -0.045 at 1.3, 2.7, and 4.0 years), a pattern consistent with temporal accumulation rather than measurement noise.

### Component-level biomarker drift over time

For descriptive context, we examined which individual AL5 components changed most over time across cohorts (Table 4). All three clinical cohorts were more likely than the general population to show biomarker worsening, with the largest cohort-specific effects occurring in the inflammation (specifically CRP) and autonomic domains (resting heart rate). Specifically, the adjusted odds of inflammatory worsening were significantly elevated for CHD (*OR* = 6.33; 95% *CI*, 1.99-38.57), AHD (*OR* = 8.84; 95% *CI*, 2.73-54.18), and NCCI (*OR* = 4.99; 95% *CI*, 1.55-30.57). Autonomic drift toward higher resting heart rates was also significant across the CHD (*OR* = 1.39; 95% *CI*, 1.05-1.86), AHD (*OR* = 1.44; 95% *CI*, 1.07-1.97), and NCCI groups (*OR* = 1.88; 95% *CI*, 1.42-2.53). Glycemic worsening (HbA1C) reached significance only in AHD (*OR* = 1.96; 95% *CI*, 1.11-3.76); HDL and waist-to-hip ratio drift did not differ significantly across cohorts. Read as a whole, this cross-domain pattern is consistent with the broader allostatic-load literature, in which inflammatory and autonomic markers tend to shift earlier under sustained physiological demand than do metabolic or anthropometric markers.

**Table 4.** Adjusted Odds of Allostatic Load Component Worsening Relative to General Population.

| **AL5 component** | **CHD OR (95% CI)** | **AHD OR (95% CI)** | **NCCI OR (95% CI)** |
| --- | --- | --- | --- |
| **Inflammation (CRP)** | **6.33 (1.99, 38.57)** | **8.84 (2.73, 54.18)** | **4.99 (1.55, 30.57)** |
| **Heart rate** | **1.39 (1.05, 1.86)** | **1.44 (1.07, 1.97)** | **1.88 (1.42, 2.53)** |
| **Glycemia (HbA1c)** | 1.56 (0.90, 2.93) | **1.96 (1.11, 3.76)** | 1.66 (0.95, 3.14) |
| **Lipids (HDL)** | 1.34 (0.80, 2.37) | 1.38 (0.80, 2.54) | 1.21 (0.71, 2.17) |
| **Anthropometry (waist-hip)** | 0.29 (0.07, 1.56) | 0.54 (0.06, 3.71) | 0.53 (0.11, 3.06) |

*Note: Adjusted for age, sex, race, Charlson index, and inter-observation interval. Bold indicates $p < .05$ (95% CI does not cross 1.0). To prevent substitution artifacts, longitudinal worsening in the inflammation domain was strictly constrained to paired CRP measurements.*

## Discussion

This study provides the first application of the allostatic load framework to adults with congenital heart disease and confirms that this population carries a significantly elevated multi-system biomarker burden relative to a general population comparison cohort. Further, exploratory findings give that gap its clinical shape.

### The gap in allostatic load is distinctly medical

Cumulative comorbidity and cardiac treatment burden explained most of the CHD-GP difference, and within the CHD cohort, cardiac medication burden was the strongest predictor of allostatic load. This factor outranked the Charlson index, age, and anatomical complexity. The original lesion category is a poor proxy for long-term biomarker trajectory in adult CHD; the physiological toll these patients accumulate over decades of cardiac management dictates their clinical course. This finding converges with the broader ACHD clinical literature documenting how acquired comorbidities increasingly dominate outcomes as the population ages,^4,17^ and it carries a direct surveillance implication: the long-term systemic burden of adult CHD is not adequately captured by complexity-tier risk stratification alone.

The fact that AHD showed a larger adjusted gap than CHD merits brief comment, because the contrast provides important clinical context. Two interpretations are compatible with the data. First, AHD represents recent dysregulation acting on a previously intact regulatory system, and the biomarker signature has not yet been softened by adaptation. Second, CHD’s lifelong adaptation may have reshaped homeostatic set points so that peripheral biomarkers underestimate the regulatory cost of sustaining them. The prospective finding that follows, demonstrating subjective mental health predicts allostatic load change uniquely within the CHD cohort, favors the second interpretation.

### Subjective mental health prospectively tracks allostatic load in CHD

This is the most provocative finding and the one that most clearly merits replication. In CHD and NCCI, worse baseline mental health predicted faster allostatic load worsening; in GP and AHD, baseline physical health predicted this physiological decline. The two cohorts where mental health predicted biomarker change share a structural feature that distinguishes them from the other two: the underlying condition is present from birth or of long duration, and the patient’s adult physical-health baseline has been shaped by it. AHD typically involves adult-onset disease layered onto a pre-morbid physical-health baseline, and GP involves no chronic disease at all. The pattern sorts cohorts by whether developmental embedding has already remade what “physical health” feels like from the inside.

A reasonable reading is that when the biomarker snapshot underestimates regulatory strain, subjective mental state captures the unmeasured burden. This underestimation occurs because lifelong adaptation has reshaped what “normal” looks like for that body. This pattern is broadly consistent with the neurovisceral integration framework,^18,19^ which locates autonomic regulation and higher-order appraisal of demand within shared neural architecture and predicts that mental and peripheral state should be coupled most tightly in populations with long-standing regulatory adaptation. Two converging lines of evidence support this reading. First, within-cohort variance decomposition showed that current subjective health contributes less to current allostatic load variance in CHD than in the general population (6.2% vs 10.6%), yet baseline mental health still carries forward predictive information about change. This suggests the relevant signal reflects a longer-arc coupling in which subjective state indexes regulatory reserve. Second, component-level drift analyses showed that the domains shifting most over time across all clinical cohorts were the inflammatory and autonomic ones, the two domains the neurovisceral framework predicts as proximal to central regulation. We offer this as descriptive support acknowledging the methodological limits of the inflammatory component.

The magnitude of the prospective effect is modest at the individual level (about 0.028 additional allostatic load units across 2.7 years per 1-SD lower Mental T-score) but non-trivial at population scale, particularly given that elevated allostatic load is itself an established predictor of cardiovascular and all-cause mortality across populations.^20^

### Clinical implications

The integration of psychological care into ACHD has been advocated on the basis of quality of life, adherence, and the symptom burden of mental health conditions in their own right.^4,17^ The exploratory prospective finding here adds a specifically physiological rationale: in CHD, subjective mental state appears to track future allostatic load trajectories. This positions psychological care as a potentially physiologically consequential intervention with plausible downstream effects on inflammation, autonomic balance, and metabolic burden.

Prospective trials of integrated behavioral cardiology in ACHD are now well positioned to test this hypothesis.

### Limitations

Several limitations warrant consideration, though some suggest our estimates may actually understate the true clinical reality. The *All of Us* program is a volunteer cohort with known under-representation of the most physically and socioeconomically vulnerable patients. Because CHD patients enter the cohort through a process that selects for those well enough to consent and engage with research, our sample may reflect a “healthy survivor” bias.

A critical methodological limitation involves differential data missingness and its impact on final cohort balance. While our initial cohort construction utilized exact-sex and nearest-neighbor age matching, the strict requirement for complete survey and biomarker data resulted in asymmetric attrition. Specifically, the surviving General Population comparison group was older and exhibited a different racial composition than the CHD cohort. In volunteer EHR databases, comprehensive biomarker panels are rarely collected randomly; they frequently indicate older age or higher baseline healthcare engagement. Consequently, our General Population benchmark represents an older, more medically engaged subset. This bias naturally inflates the baseline allostatic load of the comparison group. Taken together with the healthy survivor bias in the CHD cohort, this suggests that the significant CHD-GP gap we observed is a conservative underestimate of the true physiological disparity. Furthermore, the “general population” comparison cohort is defined merely by the absence of qualifying diagnostic codes in the electronic health record. It represents a cohort of “participants without documented cardiac or chronic disease,” a distinction which may slightly inflate the apparent gap compared to a true community sample.

Regarding longitudinal findings, the prospective analysis required participants to have a second biomarker panel in their chart, which inherently carries some selection bias. While baseline characteristics of the prospective subsample closely tracked the full sample, providing reassurance against gross selection bias, subtle selection on unmeasured factors remains possible. Additionally, PROMIS was assessed only once at baseline. While our sensitivity analyses argue against the strongest alternative explanations (such as regression to the mean or noise dilution), a formal reverse-direction test requiring dual-timepoint subjective health data remains necessary.

Finally, AL5 is an abbreviated allostatic load operationalization. It does not capture neuroendocrine markers (such as cortisol or DHEA), and the inflammatory component relies on a pre-specified white blood cell count fallback when CRP is unavailable. This fallback limits the inferential weight of any single-component finding involving the inflammation domain. The AL5 score as constructed represents one tractable, guideline-aligned index among several possible operationalizations.^11^

### Strengths

This study represents the first application of the well-established allostatic load framework to the adult CHD population, executed within one of the largest single-cohort biomarker datasets to date featuring paired psychometric data. The pre-specification of a single confirmatory test, alongside exploratory triangulation against AHD and NCCI cohorts, reduces the risk of inferential overreach. Strict congenital coding with systematic exclusion of chromosomal syndromes isolates the physiological footprint of structural heart disease from multisystem genetic conditions. Finally, variance-inflation diagnostics and progressive decomposition directly address the collinearity concerns that frequently complicate multi-predictor allostatic load analyses.

## Conclusion

McEwen originally defined allostatic load as the cumulative price the body pays to maintain stability through chronic demand.^7^ Our findings demonstrate that for adults with CHD, this biological price is exacted by the decades-long accumulation of medical and treatment burden. This physiological toll reflects the continuous cost of surviving. Furthermore, the finding that worse baseline mental health prospectively predicts faster allostatic load accumulation in CHD aligns with the foundational premise that the brain is the central organ of allostasis, translating subjective distress into systemic regulatory strain. This pattern is absent in adult-onset acquired heart disease, suggesting that lifelong cardiac survivorship reshapes the coupling between subjective state and physiological reserve. If these findings replicate, they support a clinical posture for adult CHD that treats acquired multimorbidity and mental health as fundamental drivers of long-term allostatic load.

## Data Availability

The data used in this study are available through the All of Us Researcher Workbench (https://www.researchallofus.org/). To protect participant privacy and comply with the program's informed consent, the individual-level data are not publicly available. Access to the data requires researchers to register with the All of Us Research Program, complete mandatory responsible conduct of research training, and be affiliated with an institution that has signed a Data Use Agreement. Once approved, researchers can access the Controlled Tier dataset used in this analysis via the cloud-based Researcher Workbench platform.

## Acknowledgments

The All of Us Research Program is supported by the National Institutes of Health, Office of the Director. We also thank the participants and everyone contributing to the collection of these data.

## Graphical Abstract. Visual Summary of Allostatic Load in Adults with Congenital Heart Disease

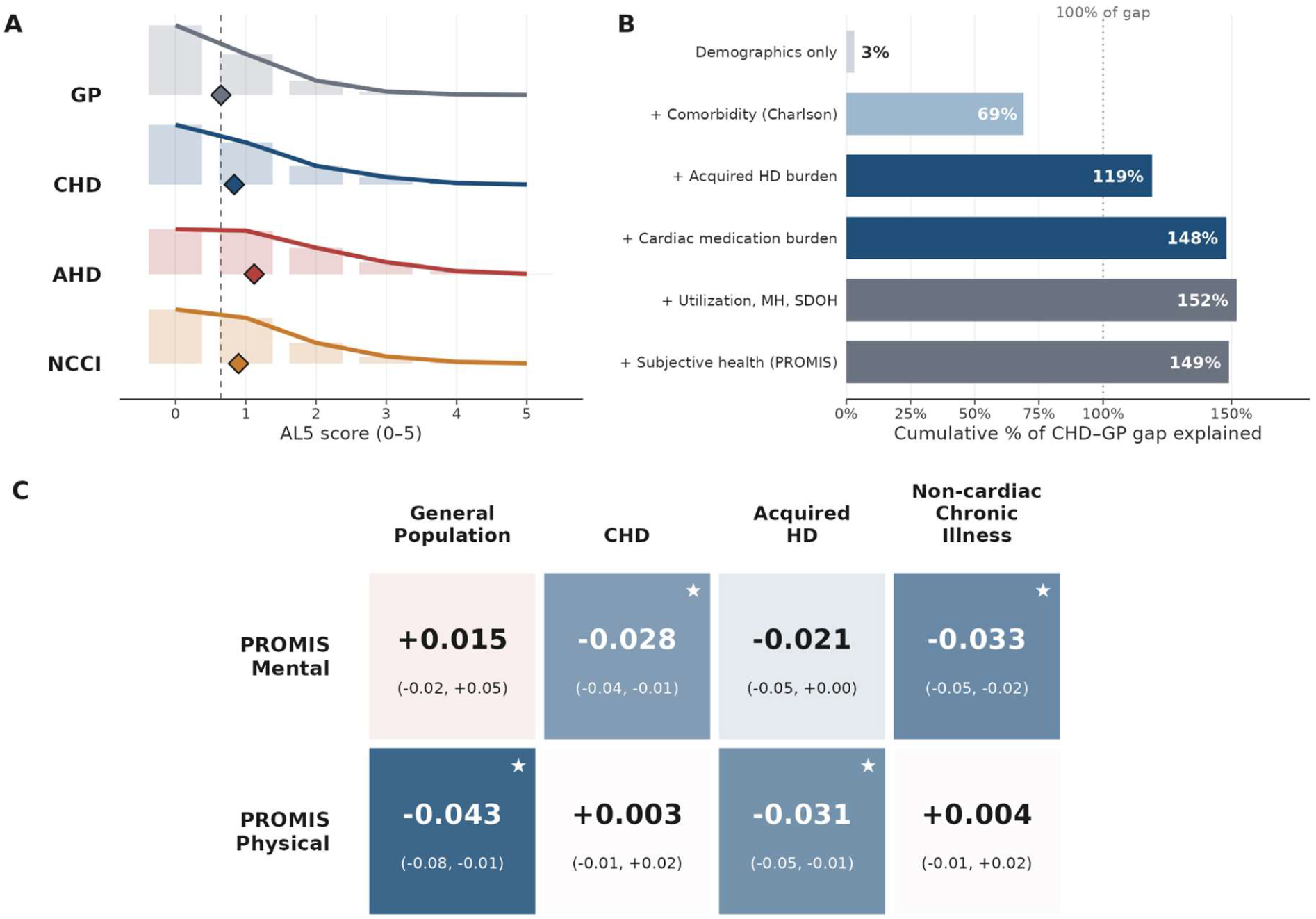

**Legend:** A three-panel composite illustrating the magnitude, drivers, and prospective implications of allostatic load (AL5) in adult CHD. **A**. *The Gap:* Adjusted cross-sectional AL5 distributions by cohort, demonstrating a significantly elevated biological burden in the CHD cohort (+0.30 AL5 units) compared to the general population (GP) reference. **B**. *The Drivers:* Progressive variance decomposition of the CHD-GP gap. The original anatomical defect does not drive the elevated allostatic load; rather, the gap is entirely explained by the accumulation of acquired medical burden, notably the Charlson Comorbidity Index (explaining 69% of the gap) and cardiac medication intensity. **C**. *Clinical Impact (Table 5 visual summary):* Cohort-specific dissociation of subjective health as a prospective predictor of allostatic load accumulation. Cell color represents the slope of AL5 change over time per +1 SD higher baseline PROMIS T-score (Blue = better baseline subjective health predicts less physiological wear; Red = positive slope; Gray = non-significant). Stars (★) indicate 95% confidence intervals excluding zero. In lifelong CHD survivorship, subjective mental health, rather than physical health, uniquely tracks the future depletion of physiological reserve. This pattern is notably absent in adult-onset acquired heart disease.

